# Unpredictable chaos or an individual stable habit – a correlational analysis of repeatedly assessed tooth brushing behavior

**DOI:** 10.1101/2023.12.26.23300550

**Authors:** Ulrike Weik, Thorben Sämann, Zdenka Eidenardt, Sadhvi Shankar Subramanian, Bernd Wöstmann, Jutta Margraf-Stiksrud, Renate Deinzer

## Abstract

Effective tooth brushing is an important part of maintaining good oral health. Epidemiological data indicate that people have difficulties to achieve oral cleanliness when brushing their teeth. Various cross-sectional studies therefore observed people while brushing and found some widespread behavioral deficits like neglect of inner surfaces and inconsistent brushing movements. Yet, longitudinal studies are missing that explore, whether people show these deficits consistently over time. To clarify this, the intra-individual stability of tooth brushing performance was investigated by repeated observation of n = 105 students at two brushing appointments (T1/T2) two weeks apart. Half of them (n = 52) were instructed to brush to the best of their ability and the other half to brush as usual (n = 53). Calibrated observers analyzed brushing behavior with respect to brushing duration, brushing movements (horizontal, circular, vertical), brushing time on tooth surfaces (outer, inner, occlusal), and distribution of time across sextants. Correlational analyses revealed a high intra-individual stability of all parameters under both instructions. Correlation coefficients varied between r = 0.72 (horizontal movements at outer surfaces) and r = 0.93 (total tooth contact time). Results indicate that people develop very specific individual toothbrushing patterns. It is important that preventive measures take into account the challenge of changing such established habits. Healthcare professionals and patients alike need to recognize this challenge when addressing oral hygiene deficiencies.

## Introduction

Thorough oral hygiene is considered a preventive self-care behavior for maintaining good oral health [1–4]. However, the high prevalence of plaque-associated periodontal disease [5–7] suggests that individuals are not capable of effectively ensuring oral cleanliness by their oral hygiene behavior. In line with this, several studies consistently showed remaining plaque immediately after tooth brushing, particularly on the sections of the gingival margin and on inner teeth surfaces [8–18] Video-based observational studies indicate that the inadequate plaque removal might be a result suboptimal oral hygiene skills [13, 15, 18–22]. In these studies, brushing performance is assessed by the analysis of brushing time and its distribution across teeth surfaces and sextants as well as of brushing movements. Results of these studies have shown that many study participants leave out at least one sextant and neglected the inner teeth surfaces, some of them did not brush inner surfaces at all. With respect to brushing techniques, study participants frequently show horizontal brushing movements instead of applying more elaborated brushing movements like circular or vertical movements [18–22].

However, to date, these observational studies are of cross sectional nature, and therefore provide limited insight into whether once-observed tooth brushing behavior is stable over time. Survey studies indicate that most people perform tooth-brushing regularly as a part of daily routine [23–25] which presumably renders it to become a habit. Habits are established as a result of a repeated and patterned sequence of behavior. Over time this behavior becomes increasingly routine and automatic [26–28]. With regard to tooth brushing behavior, children learn to brush their teeth at a very early age, even if initially with adult support and supervision [29]. Over time a tooth brushing habit is formed as a generally routinized behavior that is performed automatically and more or less subconsciously [26, 28]. This suggests that once a tooth brushing habit is established, it tends to remain stable over time. However, there is a paucity of empirical work on habit formation most of which focuses on specific aspects such as the cues that trigger the brushing behavior [30], the frequency of brushing or the sequence of actions to be performed [26]

In terms of tooth brushing performance itself, there is some data on the stability of specific aspects of the performance such as brushing duration or the brushing force. For example, it has been shown that children or students show little variation in brushing time, brushing force or other brushing patterns such as the surfaces of the teeth brushed or the brushing technique [31–33]. However, these studies date back a long time or the data are based only on a small subsample of the study participants. Furthermore, in most cases only a few aspects of the tooth brushing behavior such as brushing duration or brushing force were investigated. Thus, it still remains uncertain, which further parts of the brushing performance itself are displayed in a stable form or vary randomly from brushing to brushing event. This knowledge is important when it comes to alter aspects of brushing behavior in order to improve its effectivity. The aim of the present study is therefore to examine whether the tooth brushing parameters analyzed in the above-mentioned observational studies occur in the same or similar extent when the brushing performance of individuals is not only observed once but repeatedly over time. In some of the mentioned observational studies participants were instructed to brush their teeth “to the best of their ability” [15, 17, 21, 22] and in others to “brush as usual” [18]. The present study examines the stability of the brushing behavior in terms of both best brushing and brushing as usual. It is hypothesized that, irrespective of best or usual brushing the repeatedly observed behavioral parameters will be highly stable.

## Methods

### Ethics

The study was conducted according to the principles of the Declaration of Helsinki. The Ethics Committee of the Department of Medicine at Justus Liebig University Giessen approved the study protocol (file no 254/18; 2019/01/23). The study has been registered at the German clinical Trials Register (www.drks.de; ID: DRKS00017812; 2019).

Recruitment of study participants began 04/05/2019 and ended 07/17/2019. Study participants received detailed information before the start of the examination and all participants gave their written consent. The study had two objectives: the comparison of brushing as usual and brushing to the best of one’s abilities (data already published in 34) and the analysis of the stability of different parameters of tooth brushing behavior. The data presented here refer to the second objective, i.e. the stability of behavioral parameters observed at two time points (T1 and T2) within an interval of two weeks.

### Study sample

Study participants were university students from Giessen (Hesse, Germany) aged between 18 and 35 years, who predominantly (at least 2/3 of all brushing events) brush their teeth with a manual toothbrush. Participants were recruited through the university’s internal email distribution list, which covers almost all students at Justus Liebig University, and through advertisements in a regional online magazine. Subjects were excluded from study participation if any of the following criteria applied: (1) study of dentistry or human medicine, (2) fixed orthodontic appliances, removable prostheses/dentures, dental jewelry or oral piercings, (3) physical impairment affecting oral hygiene behavior, (4) use of antibiotics within the last three months prior to study entry, (5) dental prophylaxis within the last four months prior to study entry, and (6) pregnancy. Sample size calculation using the free available power analysis program G*Power [35] was based on the first study objective (comparison of brushing to the best of one’s ability vs. as usual) and resulted in a minimal sample size of n = 102 (see Weik et al., 2023). With regard to the present study, this sample size allows for the detection of correlations of ρ > 0.72 with alpha = 0.05 and a power of 1 - β = 0.80.

### Procedures

The study was conducted at the Institute of Medical Psychology of the Justus Liebig University in Giessen. Procedures are already described in detail in 34) and provided in the supplemental material. Briefly, after being informed about the study, students who were eligible to participate were scheduled for two brushing sessions two weeks apart. Procedures at the two appointment (T1 and T2) were the same. While participants brushed their teeth at T1 and T2, clinical data were not assessed on both appointments. Disclosing of teeth and assessment of dental plaque took place only at T2. To keep both appointments as similar as possible and to prevent visible plaque staining at T1 from influencing brushing behavior at T2, a sham staining (with water faked as a fluorescent solution) and a simulated plaque assessment was carried out at T1. With regard to the first study objective (not focused here) study participants were randomly assigned to two brushing conditions with different brushing instructions (brushing to the best of one’s abilities vs. brushing as usual). At both appointments study participants brushed their teeth according to their assigned brushing instructions. Stuff persons involved in data assessment were not changed between the two appointments and were blinded to the respective brushing condition. Any Interaction with study participants was conducted in a fully standardized manner. At both brushing sessions study participants were placed in front of mobile wash basin and a tablet with a front camera mounted at a tripod. The tablet served as a mirror for the participants as well as a device for recording the tooth brushing performance. In addition to the tablet, tooth brushing was also recorded by two side cameras mounted on the walls in case the tablet camera did not fully capture the brushing event. The participants were provided with a standard manual toothbrush (Elmex InterX short brush-head, medium; CP GABA, Hamburb, Germany) and toothpaste (Elmex; CP GABA, Hamburg, Germany). Additionally, dental floss (waxed and unwaxed dental floss; Elmex; CP GABA, Hamburg, Germany), super floss (Meridol Special-Floss; CP GABA, Hamburg, Germany) and interdental brushes (Elmex interdental brush sizes 2 and 4; CP GABA, Hamburg, Germany) were provided on a table beneath the basin. After they received the respective brushing instruction they were asked to start brushing their teeth and their brushing performance was videotaped.

### Observed behavioral parameters

The video-based analysis of the tooth brushing behavioral parameters was conducted according to the procedure described in previous studies [17, 21]. A detailed description is provided in the supplemental material. The videos were analyzed by the use of an observational software (Interact 18; Mangold International; Arnsdorf, Germany). Videos were analyzed by independent calibrated observer. For calibration five videos from a previous study were used. Criterion for successful calibration were intra-class correlations (ICC) ≥ 0.90 for five consecutively analyzed videos for each of the observed behavioral parameters. In order to ensure reliability of video analysis an additional 10 videos of the study participants were analyzed in duplicate by two independent observers. Determination of the ICC of these double codings resulted in a high agreement between the respective independent observers (ICC ≥ 0.90 for all observed behavioral parameters). Behavioral parameters were the following: a)Total tooth contact time (tct; time (seconds) in which the brush touches the teeth); b) tct and proportional distribution of tct on the tooth surfaces (occlusal, outer and inner teeth surfaces, respectively); c) tct spent with circular or horizontal brushing movements on outer surfaces and vertical and horizontal movements on inner surfaces; d) overall quality index for the distribution of tct across sextants and teeth surfaces (QIT-S; Deinzer et al., 2018; this index represents a rank-scaled measure describing the extent to which the sextants were brushed on outer and inner surfaces, respectively).

### Statistical analysis

The present data analysis was conducted to test the hypothesis that the described tooth brushing parameters observed at T1 and T2, will be highly correlated – regardless whether study participants brushed to the best of one’s ability or as usual. According to [36] correlational coefficients between 0.70 and 0.90 indicate a high relationship, correlations above 0.90 describe a very high correlation and a very dependable relationship.

A re-test correlation of r > 0.70 is regarded high enough to consider tooth brushing behavior as a stable behavior over time. In addition to the product-moment correlational coefficient (Pearson) rank correlation coefficients (Spearman) were calculated in order to account for potential outlier values. According to the research hypothesis the following hypothesis pair was statically tested independently for each of the behavioral parameters under observation: H_0_: ρ_T1/T2_ ≤ 0.5; H_1_: ρ_T1/T2_ > 0.5. For description of these brushing parameters the respective means and SDs and effect sizes with 95% confidence intervals with respect to differences of the means at T1 and T2 are reported. Effect sizes are calculated for correlational data according to Dunlap [37]. According to Cohen [38], effect sizes of d ≥|.2| |.5| |.8| are considered small, medium and large, respectively. The relationships between T1 and T2 of the categorical data assessed by the QIT-S index were analyzed by chi^2^-tests. All statistical analyses were conducted with the statistical software package SPSS (IBM SPSS Statistics for Windows, Version 28, IBM, Armonk, New York, USA).

## Results

106 study participants finished the study. One person in the best-brusher group was excluded from analyses due to a very unusual brushing behavior (tooth contact time exceeded 15 minutes and deviated from the mean value by more than four SD). Description of the study sample with respect to demographic and clinical data is given in Table 1. Descriptive data and results of correlational analyses of the observed brushing parameters are shown in Table 2 for brushing to the best of one’s ability and Table 3 for brushing as usual. All reported correlational coefficients were statistically significant (all p > 0.001). Scatterplots for each behavioral parameter are shown in the supplemental material (figures in S1). Cross tables in Fig 1 illustrate the relationship of the QIT-S scores at T1 and T2 showing high concordance for both, best brushers (outer surfaces: chi^2^ = 16.65, p < 0.001; inner surfaces: chi^2^ = 146.92, p < 0.001) and as usual brushers (outer surfaces: chi^2^= 30.23, p < 0.001; inner surfaces: chi^2^ = 114.01, p = 0.001).

**Fig 1.**
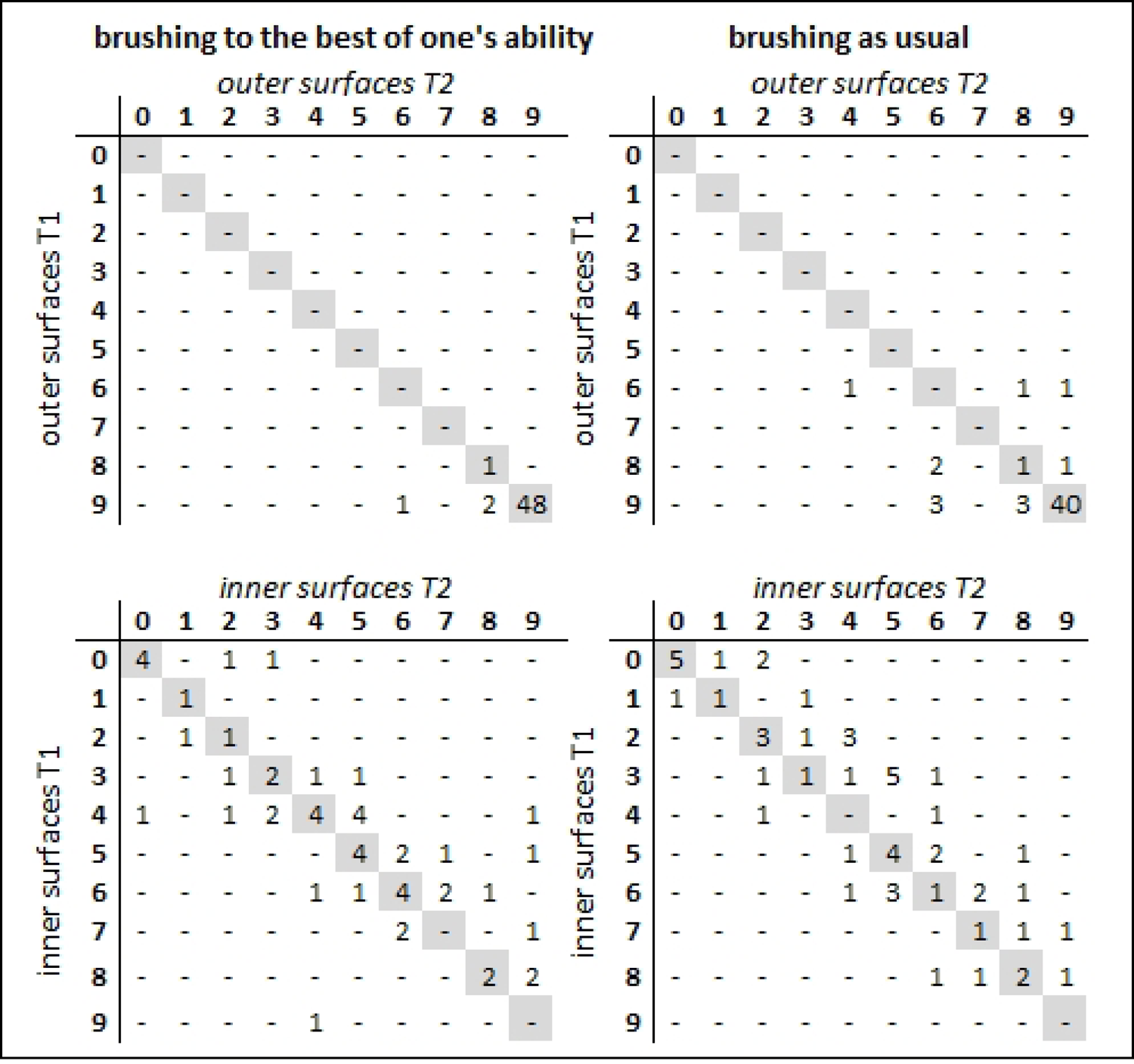
Relationship of QIT-S scores between T1 and T2. Cross tables on the left show the QIT-S for brushing to the best of one’s ability for outer and inner surfaces respectively; cross tables on the right show the QIT-S for brushing as usual for outer and inner teeth surfaces, respectively. QIT-S scores: 0–5: 0–5 sextants were brushed for at least one second; score 6: every sextant was brushed for at least 1 s but less than 3.5 s; score 7 and 8: every sextants was brushed between 3.5–5 s and 5–7.5 s, respectively; score 9: all sextants were brushed for at least 7.5 s.

**Table 1.** Description of the study sample.

| Table 1. Description of the study sample |  |  |  |  |
| --- | --- | --- | --- | --- |
|  | Brushing to the best of one's ability (n=52) |  | Brushing as usual (n=53) |  |
|  | M (SD)<br>n/n |  |  |  |
| Sex (f/m/x) | 42/9/1 |  | 45/8/0 |  |
| Age (years) | 22.77 (2.5) |  | 23.49 (3.0) |  |
| DMFT-index | 2.77 (3.4) |  | 3.74 (3.6) |  |
|  | T1 | T2 | T1 | T2 |
| PBI mean | 0.73 (0.4) | 0.72 (0.4) | 0.64 (0.4) | 0.66 (0.4) |
| PBI % bleeding full mouth | 43.41 (17.6) | 43.22 (18.7) | 38.72 (19.0) | 41.02 (20.1) |

DMFT: decayed, missed, filled teeth; PBI: papillary bleeding index; T1: first brushing session; T2: second brushing session

**Table 2:** Results of behavioral parameters (brushing to the *best of one’s ability*) at T1 and T2 (n=52)

| *Tooth brushing parameters* | T1 |  | T2 |  | *d* | [CI 95%] | *r* * | [CI 95%] | *rho* * | [CI 95%] |
| --- | --- | --- | --- | --- | --- | --- | --- | --- | --- | --- |
|  | *M* | *SD* | *M* | *SD* |  |  |  |  |  |  |
| Tooth contact time (tct; s) | 234.06 | 90.15 | 260.02 | 118.73 | -0.21 | -0.34 -0.08 | 0.90 | 0.84 0.94 | 0.85 | 0.74 0.91 |
| Occlusal surfaces (s) | 89.11 | 48.01 | 100.23 | 50.09 | -0.23 | -0.39 -0.06 | 0.84 | 0.74 0.91 | 0.77 | 0.63 0.87 |
| Outer surfaces (s) | 106.35 | 48.10 | 111.54 | 54.09 | -0.10 | -0.23 0.04 | 0.89 | 0.81 0.93 | 0.84 | 0.73 0.91 |
| Inner surfaces (s) | 38.61 | 32.64 | 48.25 | 47.10 | -0.21 | -0.38 -0.04 | 0.83 | 0.72 0.90 | 0.84 | 0.74 0.91 |
| Occlusal surfaces (%tct) | 37.96 | 14.36 | 39.07 | 12.89 | -0.08 | -0.25 0.08 | 0.83 | 0.71 0.90 | 0.79 | 0.65 0.87 |
| Outer surfaces (%tct) | 46.51 | 12.59 | 44.41 | 13.84 | 0.16 | -0.02 0.34 | 0.80 | 0.67 0.88 | 0.81 | 0.68 0.89 |
| Inner surfaces (%tct) | 15.54 | 10.78 | 16.51 | 11.49 | -0.09 | -0.28 0.10 | 0.77 | 0.63 0.86 | 0.78 | 0.64 0.87 |
| Outer surfaces |  |  |  |  |  |  |  |  |  |  |
| Horizontal movements (s) | 31.80 | 31.81 | 34.10 | 39.91 | -0.06 | -0.27 0.15 | 0.72 | 0.55 0.83 | 0.80 | 0.66 0.88 |
| Circular movements (s) | 68.02 | 51.14 | 69.05 | 51.09 | -0.02 | -0.16 0.12 | 0.87 | 0.79 0.92 | 0.82 | 0.69 0.89 |
| Inner surfaces |  |  |  |  |  |  |  |  |  |  |
| Horizontal movements (s) | 18.32 | 18.78 | 22.66 | 25.41 | -0.18 | -0.36 0.00 | 0.80 | 0.67 0.88 | 0.80 | 0.67 0.88 |
| Vertical movements | 15.51 | 23.67 | 19.14 | 33.37 | -0.11 | -0.28 0.06 | 0.81 | 0.70 0.89 | 0.67 | 0.49 0.80 |

T1: first brushing session; T2: second brushing session; tct: tooth contact time; s: seconds; d: effect size Cohen's d for differences of means; CI 95 %: 95% confidence interval; \*all p < 0.001.

**Table 3:**
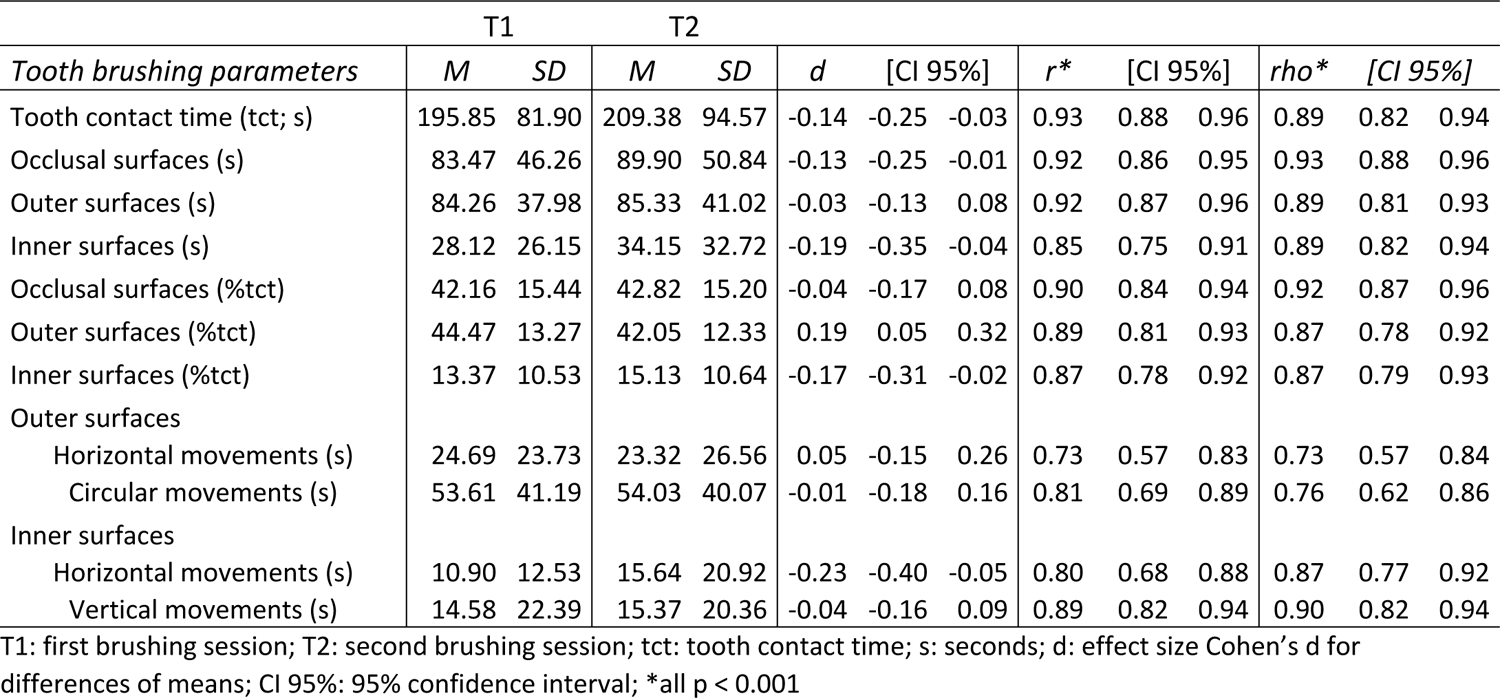
Results of behavioral parameters (brushing *as usual*) at T1 and T2 (n=53)

## Discussion

Previous studies observing tooth brushing behavior have been of cross-sectional nature so far. The aim of the present study was to examine whether various aspects of this behavior would be stable over time. To achieve this, the study participants brushed their teeth twice at two brushing session two weeks apart. Half of the study sample brushed their teeth to the best of their ability, and the other half brushed as usual. As tooth brushing behavior is considered to be a rather routine and automated behavior [26, 28], it was expected that the performance of tooth brushing would be very similar at both time points (T1 and T2), i.e. that a person’s position in the group would remain fairly stable over time.

The results support this assumption as the null hypothesis (H_0_: ρ_BV1,MT_ ≤ 0.5) is rejected for all behavioral parameters. As expected, correlational analyses revealed high correlations for the observed behavioral parameters. In particular, the observed absolute time values show strong correlations (≥0.80) indicating a stable behavior. Even though the “best” brushers showed a longer brushing time (i. e. tooth contact time (tct)) than the “as usual” brushers correlations of ≥ 0.90 were found in both groups indicating a high degree of concordance between T1 and T2 in individuals, regardless of whether they brushed to the best of their ability or as usual. The fact that in both groups the brushing times were somewhat longer at T2 compared to T1 (see table 2 and 3) could be interpreted as a carry-over effect from the procedures of the first brushing session. At T1 a plaque measurement was simulated by (sham-) staining after tooth brushing. Although this sham procedure did not give the subjects any realistic feedback about plaque on their teeth, it may have encouraged them to make a greater effort to clean their teeth at T2, and thus to increase their brushing time. However, as the correlational analyses show this potential increase in brushing time is evident for all individuals equally.

Similar to the total brushing time, the absolute time values for brushing on occlusal, outer and inner surfaces, respectively, are highly stable over time. In addition, the proportional distribution of time to the teeth surfaces shows a close relationship between T1 and T2. In fact, the proportion of brushing time spent on the respective tooth surfaces found here are similar to those already observed in students or adults with only minor variations [16, 17, 19]. These results suggest that the way individuals brush their teeth seems to be a rather stable behavior, which is even more evident when considering the brushing technique, i. e. brushing movements used by the study participants. It was already reported many years ago, that among different aspects of tooth brushing the applied brushing technique seems to be the one with the most pronounced habit when repeatedly observed [32]. With respect to the brushing technique previous observations showed that the majority apply circular and horizontal movements on the outer tooth surfaces and vertical and horizontal movements on inner surfaces [15, 17, 21, 22, 34]. Therefore, the present analyses focused on these brushing movements. Although somewhat less pronounced than in the other brushing parameters, there is also a clear relationship between T1 and T2.

Finally, the results for the QIT-S also show a concordance between T1 and T2. These results differ slightly depending on whether the outer or inner surfaces are considered and also depending on whether the study participants brushed their teeth to the best of their ability or as usual. On the outer surfaces about more than 95 % of the “best” brushers brushed all sextants at least for more than 5.5 seconds, the majority even longer and only less than 5 % showed small variations. In most of the “as usual” brushers (about 75 %) there is also a strong relationship in the QIT-S for outer surfaces, although it is slightly less pronounced compared to the “best” brushers. However, the variation shown in some study participants is still rather small. The results of the QIT-S for inner surfaces are somewhat different, regardless of how the participants brushed their teeth. In contrast to the outer surfaces the sextants on the inner surfaces are brushed for much less time and some people did not brush the inner surfaces at all. This neglect of the inner surfaces has been observed in several observational studies [8, 15, 17, 20, 21, 34, 39, 40] and this neglect could be the reason for much more variation in this aspect of the brushing behavior. A strong behavioral habit for brushing inner surfaces seems to be not yet established. Nevertheless, most of the participants show values scattered around the diagonal representing a complete concordance, which was found in about 40 % of the “best” and about 30 % of the “as usual” brushers.

In summary, the data from the present study show that the tooth brushing behavior exhibited during the two brushing sessions appears to be an intra-individual stable behavior. This is consistent with the assumption that tooth brushing behavior is indeed a habit [26, 28], which is hardly surprising. Most people perform this behavior regularly on a daily basis and usually from a very early age on. Over time, it becomes a routine behavior that can be performed largely automatically. So far, research on habit formation in the context of tooth brushing behavior focused aspects such as when or whether this behavior occurs [26]. In contrast, how individuals perform this behavior and whether this performance represents a habitual pattern has received little attention. The results of the present study show that the performance of tooth brushing behavior appears to be a routine with only little variation. Consistent with this are findings of intra-individual tooth brushing patterns observed in a cross-over study, where students brushed their teeth with a manual and a powered tooth brush [20]. In this study there was no time interval between repeatedly observed tooth brushing, but even when brushing with a different brushing device, study participants tended to show a similar behavioral pattern [20].The present study has some strengths but also some limitations that should be discussed. One of the strengths is the systematic analysis of various aspects of tooth brushing behavior, such as total brushing time, its distribution on tooth surfaces and sextants and brushing technique. This, rather than looking at one or a few aspects, provides a more thorough understanding of the behavior. Still, there other important aspects of tooth brushing behavior which have not been addressed here. These include the brushing force or the sequence of brush positions and the number of times brushing areas are changed. However, there are reports that these aspects also show a considerable concordance between different brushing sessions [32, 33]. The thorough training and calibration of the video observer and the high standardization in the conduction of the study are also strengths to be highlighted. The study procedures at T1 and T2 were kept as constant as possible in order to ensure that participants’ toothbrushing was performed under the same conditions. With regard to the first study objective, plaque assessment took place at T2 before and after tooth brushing. In order to avoid a visual feedback of residual plaque at T1 and thus a change in brushing behavior, a sham staining procedure was performed at T1. However, it cannot be excluded that this sham procedure caused a change in the behaviour. The longer brushing times at T2 compared to T1 could be a result of this. Another limitation arises in terms of the external validity of the results. Firstly, the present study analysed university students. The generalizability to other age groups is yet unproven. However, tooth brushing routines are acquired through regular and frequent practice. One can thus assume that older persons show an even more pronounced habit.

However, it would be interesting to study children in this regard. Secondly, the stability of this behavior was only demonstrated in laboratory conditions and it is unclear whether individuals exhibit the same level of stability when brushing their teeth in daily life. Further studies analyzing tooth brushing in a domestic setting would be instructive.

## Conclusion

Findings suggest that people develop a stable individual pattern of toothbrushing performance – including its deficiencies. Such a stable behavior is much harder to change than a brushing behavior that would only be dysfunctional by chance. These results indicate that simply asking people to increase the frequency of toothbrushing might not improve their oral hygiene. Preventive measures should consider the difficulty of changing established habits. Healthcare professionals and patients alike need to recognize this challenge addressing oral hygiene deficiencies.

## Data Availability

All relevant data are available at: https://zenodo.org/records/10377716

Supporting information S1: File (PDF)

